# PROSPECTIVE COMPARISON OF OPTIMIZED COMPRESSED SENSING AND GRAPPA-ACCELERATED 3D-TOF-MRA FOR EVALUATION OF UNTREATED INTRACRANIAL ANEURYSMS AT 1.5 T: A NON-INFERIORITY STUDY

**DOI:** 10.64898/2026.11.02.26367077

**Authors:** Catalin George Iacoban, Holger Bösenberg, Claudia Klüner, Olga Alykova, Maike Gerdes, Franziska Meinert, Benedikt Sundermann, Christian Mathys

**Affiliations:** Institute of Radiology and Neuroradiology, Evangelisches Krankenhaus Oldenburg, Universitätsmedizin Oldenburg, Steinweg 13-17, 26122 Oldenburg, Niedersachsen, Germany; Research Center Neurosensory Science, Carl von Ossietzky Universität Oldenburg, Ammerländer Heerstraße 114-118, 26129, Oldenburg, Niedersachsen, Germany; Clinic of Radiology, University of Münster, Faculty of Medicine, Schlossplatz 2, 48149, Münster, Nordrhein-Westfalen, Germany; University Clinic of Neurosurgery, Evangelisches Krankenhaus Oldenburg, Universitätsmedizin Oldenburg, Steinweg 13-17, 26122 Oldenburg, Niedersachsen, Germany; Department of Neuroradiology, Istituto Giannina Gaslini, via Gerolamo Gaslini 5, 16147 Genova, Liguria, Italy

## Abstract

**RATIONALE AND OBJECTIVES:** 3D-TOF-MRA is a standard non-invasive method for detecting and monitoring intracranial aneurysms, but long scan times remain a limitation. Compressed sensing (CS) acceleration for intracranial aneurysm evaluation has mainly been studied at 3T. This prospective single-center non-inferiority study compared CS-accelerated 3D-TOF at 1.5T with GRAPPA-accelerated 3D-TOF-MRA for evaluating untreated intracranial aneurysms.

**MATERIALS AND METHODS:** Sample size was determined via a preliminary retrospective study. 25 patients with 32 untreated aneurysms underwent both CS- (factor 7, 3:04 min) and GRAPPA-accelerated (factor 2, 5:00 min) 3D-TOF, yielding a 38.7% scan-time reduction. Two blinded neuroradiologists assessed overall image quality (primary endpoint) and artifacts, subjective CNR, aneurysm delineation and diameters (secondary endpoints). Non-inferiority was tested via one-sided t-test (prespecified 10% margin); interrater agreement used ICC.

**RESULTS:** CS demonstrated non-inferiority to GRAPPA for overall image quality in both readers (p ≤ .023), though interrater agreement was low (ICC 0.241–0.262), reflecting subjective variability. Secondary exploratory analyses showed heterogeneous preferences. ROI-based CNR was lower for CS (median 169.6; IQR, 91.0-226. vs. 278.2; 181.0-366.2). Aneurysm diameter measurements showed excellent interrater agreement for both techniques (CS ICC, 0.899-0.980; GRAPPA ICC, 0.877-0.973; all p < .001).

**CONCLUSION:** CS-accelerated 3D-TOF at 1.5T met non-inferiority criteria vs. GRAPPA-TOF for overall image quality, reducing scan time by 38.7% while preserving aneurysm measurement reliability. However, reduced CNR and reader-preference variability important trade-offs.

These findings support the use of CS-accelerated-TOF as a time-efficient alternative to GRAPPA-accelerated-TOF in evaluating intracranial aneurysms, though further research on field-strength effects and small aneurysms is warranted.

## INTRODUCTION

Long scan times and motion artifacts continue to limit MR performance in clinical practice. Shorter acquisitions improve patient comfort, reduce motion artifacts, and enhance efficiency (1,2). With newer scanner generations, advanced acceleration techniques make this increasingly feasible in routine imaging (3,4).

Parallel imaging (PI) techniques such as GRAPPA (Generalized Autocalibrating Partially Parallel Acquisition) and CAIPIRINHA (Controlled Aliasing in Parallel Imaging Results in Higher Acceleration) substantially shorten acquisition times across many MR sequences (5–9).

More recently, compressed sensing (CS) has emerged as an additional acceleration method, exploiting the inherent sparsity of MR data, enabling undersampled acquisitions and reconstruction of full datasets for shorter scan times or higher spatial resolution (10).

However, acceleration is constrained by potential image quality loss and reconstruction-related artifacts (11–14).

TOF-MRA is an established non-invasive technique for the detection and follow-up of intracranial aneurysms (15,16). Owing to its high spatial resolution and ability to visualize intracranial vessels without contrast administration or ionizing radiation, TOF-MRA is widely used for screening and surveillance, particularly in patients requiring repeated imaging.

The advantages of CS in TOF-MRA have been demonstrated in studies of extracranial and intracranial arterial pathologies, including stenoses and moyamoya disease (17–19). These studies showed that CS-based TOF-MRA reduces acquisition time while maintaining or improving image quality, vessel delineation, and artifact reduction compared with conventional TOF-MRA (20,21).

In aneurysm-focused studies, mainly at 3T, CS-TOF has shown comparable or improved image quality versus PI-TOF, with fewer artifacts and equivalent aneurysm size measurements when validated against DSA (22).

To date, no studies have systematically compared CS- with conventional GRAPPA-acceleration for intracranial aneurysm assessment using TOF-MRA on widely available 1.5T systems, as most prior work has focused on 3T. In this prospective paired non-inferiority sequence-comparison study, we compared optimized CS- and GRAPPA-accelerated 3D-TOF at 1.5T in clinical routine, using overall image quality as the primary endpoint and additional image quality assessments and interrater reproducibility of aneurysm diameter measurements as exploratory secondary endpoints.

## METHODS

### RESEARCH DESIGN

This single-center prospective sequence-comparison study was approved by our institutional ethics committee of Carl von Ossietzky Universität Oldenburg, Germany (registration no. 2021-033). Sample size was estimated based on a precursor pilot study with a retrospective analysis of routine clinical MRI data. Written informed consent was obtained from all participants in accordance with the Declaration of Helsinki. No contrast agent or sedative medication was administered for study purposes. However, patients who received contrast agents and/or sedation for clinical reasons were eligible.

During the first stage, sequence parameters for newer (CS) and conventional (GRAPPA) acceleration techniques were optimized in five healthy participants using a pragmatic empirical approach (see supplementary material). Based on expert consensus, the best-performing sequence from each acceleration arm was selected for the clinical phase (Table 1).

**Table 1.** Imaging parameters for the CS and GRAPPA-accelerated 3D-TOF.

| **Parameters** | **CS-3D-TOF (Sequence No. 20)** | **GRAPPA-3D-TOF (Sequence No. 19)** |
|---|---|---|
| TE/TR (ms) | 7,15 / 23,3 | 7,15 / 23,3 |
| FA (Flip Angle) | 20<sup>o</sup> | 20<sup>o</sup> |
| FOV (Frequency x Phase) (mm) | 220 x 197 | 220 × 197 |
| Nominal Resolution (mm) | 0.73 x 0.72 x 0.78 | 0.82 x 0.81 x 0.99 |
| Scan length (Z-axis) (mm) | 81,6 | 81,6 |
| Number of 3D blocks | 4 | 4 |
| Number of Slices | 136 | 136 |
| Interpolation | 1,5 | 1,5 |
| Slice Thickness (mm) | 0.6 | 0.6 |
| Matrix (Frequency x Phase) | 304 x 269 | 272 x 242 |
| Bandwidth (Hz/Pixel) | 100 | 100 |
| Acceleration factor | 7 (CS factor) | 2 (GRAPPA factor) |
| Denoising | auto | N/A |
| Scan time (min:sec) | 3:04 | 5:00 |

During the second stage, the previously optimized CS-accelerated and GRAPPA-accelerated sequences were added to routine clinical MRI examinations of patients with at least one known untreated intracranial aneurysm, as indicated by neurological/neurosurgical referral information. Both sequences were acquired natively, although the overall protocol might have included contrast administration. The acquisition order of CS and GRAPPA was systematically alternated. The subsequent comparison included an assessment of general and specific aspects of image quality as well as measurement of aneurysm diameters.

### ENDPOINTS

The primary endpoint was overall image quality, with the primary hypothesis that optimized CS-accelerated 3D-TOF would not result in a clinically relevant reduction in overall image quality (>10%) compared with GRAPPA-accelerated TOF. Secondary endpoints included comparison of general image artifacts (numerical rating scale), motion artifacts (numerical rating scale), CNR (numerical rating scale and ROI-based measurements), aneurysm delineation (numerical rating scale), overall aneurysm depiction (numerical rating scale) as well as aneurysm diameters.

### PATIENT COHORT

During sequence optimization, 5 healthy volunteers (≥18 years) were recruited. Based on the sample size estimation pilot study, the final cohort comprised 25 patients with at least one known untreated intracranial aneurysm. Between July 2024 and December 2024, 25 consecutive patients at the Institute of Radiology and Neuroradiology, Evangelisches Krankenhaus Oldenburg underwent MRI at 1.5T as clinically indicated. Standard MRI contraindications and specific CNS disorders were exclusion criteria (see supplementary material).

### IMAGING PROTOCOL

Imaging was performed on our clinical 1.5 T whole-body MR scanner (Magnetom Sola 1.5T; SIEMENS Healthineers, Erlangen, Germany; Software version XA20, 20 channel head/neck receiver coil, installed February 2020). Primary image data and technical parameters were stored in standard DICOM format.

The individualized MRI protocol included the two preselected, optimized CS- and GRAPPA-accelerated TOF-MRA sequences in addition to clinically indicated sequences.

A CS acceleration factor of 7 was applied, resulting in a scan time of 3:04 min:s. For GRAPPA-accelerated TOF, a GRAPPA factor of 2 was used, yielding a scan time of 5:00 min:s (38.7% scan time reduction). Both 3D-TOF-MRA datasets were acquired in the axial plane. Detailed imaging parameters are summarized in Table 1.

### IMAGE ANALYSIS

All 3D-TOF datasets where pseudonymized, randomized, and independently assessed by two blinded readers (7 (CGI) and 14 years’ (BS) neuroradiology experience). Readers were blinded to sequence identity, but aneurysm number and location were provided in advance for each patient. Each patient was analyzed only once per session, with either CS-accelerated or GRAPPA-accelerated 3D-TOF. Subjective image quality parameters were rated on a 100-point scale (0 = poor (insufficient for diagnosis / severe artifacts), 50 = adequate (moderate artifacts not impairing diagnosis), 100 = excellent (no artifacts)) across predefined criteria (see endpoints). CNR was calculated for each TOF acquisition using predefined ROIs in the aneurysm lumen, brain stem and background noise. Aneurysm diameters were measured in a standardized manner and recorded numerically in millimeters (mm) (see supplementary material). Mean and maximum diameters were calculated. As temporally matched DSA as reference standard was unavailable for most patients, neither diagnostic detection accuracy nor measurement accuracy relative to ground truth were assessed.

### STATISTICAL ANALYSIS

Statistical analyses were performed using SPSS Version 30 (IBM, Armonk, NY, USA) and SAS^®^ Analytics Software (Online Studio) (SAS Institute, Cary, NC, USA). Standard descriptive statistics were applied.

Non-inferiority of CS relative to GRAPPA for the primary endpoint (overall image quality) was assessed separately for each rater using log-transformed scores. This was done using a one-sided t-test with a prespecified non-inferiority margin of 10%, a difference considered clinically acceptable for overall diagnostic performance (23). Results were visualized using box-and-whisker plots. Non-inferiority was concluded if the lower bound of the one-sided 95% CI for the mean difference (CS - GRAPPA) exceeded the predefined margin. The threshold for significance was set at a *p* < 0.05.

Secondary endpoints were summarized for each variable as median (interquartile range). Visual comparisons of secondary endpoints were performed using split-violin plots to explore potential determinants of the main result (primary endpoint).

Interrater agreements between rater A and rater B for CS and GRAPPA for all endpoints were reported as single-measures intraclass correlation coefficients derived from a two-way mixed-effects model with absolute agreement.

## RESULTS

### COHORT CHARACTERISTICS

Twenty-six patients were enrolled; one was excluded because no aneurysm was identified, leaving 25 patients with 32 aneurysms for analysis. Patient and aneurysm characteristics are summarized in Table 2. Most aneurysms were located in the anterior circulation and were intradural. Median aneurysm size was 4.1 mm (IQR 2.7–5.7), with most measuring <5 mm. **Table 2**. Patient and aneurysm characteristics

**Table 2.** Patient and aneurysm characteristics.

| **Patient characteristics (n = 25)** |  | **Aneurysm characteristics (n=32)** |  |
|---|---|---|---|
| Age (years) | 60.6 ± 13.8 | **Aneurysm size\* (mm)** | 4.1 (2.7–5.7) |
| Female sex, n (%) | 19 (76) | <5 mm, n (%) | 21 (65.6) |
| Body mass index (kg/m²) | 27.4 ± 6.5 | 5–10 mm, n (%) | 9 (28.1) |
| No. of aneurysms per patient, n (%) |  | >10 mm, n (%) | 2 (6.3) |
| 1 | 18 (72) | **Topography, n (%)** |  |
| 2 | 5 (20) | Anterior circulation | 25 (78.1) |
| 3 | 2 (8) | Posterior circulation | 7 (21.9) |
|  |  | **Dural location, n (%)** |  |
|  |  | Intradural | 23 (71.9) |
|  |  | Extradural | 9 (28.1) |
|  |  | **Parent artery, n (%)** |  |
|  |  | Internal carotid artery (ICA) | 13 (40.6) |
|  |  | Middle cerebral artery (MCA) | 5 (15.6) |
|  |  | Posterior communicating artery (PCom) | 4 (12.5) |
|  |  | Anterior choroidal artery (AChA) | 3 (9.4) |
|  |  | Anterior cerebral artery (ACA) | 2 (6.3) |
|  |  | Vertebral artery (VA) | 2 (6.3) |
|  |  | Anterior communicating artery (ACom) | 1 (3.1) |
|  |  | Posterior cerebral artery (PCA) | 1 (3.1) |

\* Aneurysm size was defined as maximum diameter measured on GRAPPA sequences using measurements from rater B.

### PRIMARY ENDPOINT

Non-inferiority of CS-acceleration relative to GRAPPA-acceleration for overall image quality was demonstrated for both raters using log-transformed scores and the prespecified 10% non-inferiority margin. For Rater B, the mean difference (CS − GRAPPA) was 0.059, with a one-sided 95% CI lower bound of 0.021 (*p* < .0001), indicating non-inferiority and a numerical preference for CS. For Rater A, the mean difference was −0.062, with a lower bound of −0.097 (*p* = .023), likewise meeting the criterion for non-inferiority, despite a numerical preference for GRAPPA (Figures 1 and 2, Table 3). Interrater agreement for overall image quality was low for both acceleration methods.

**Figure 1.**
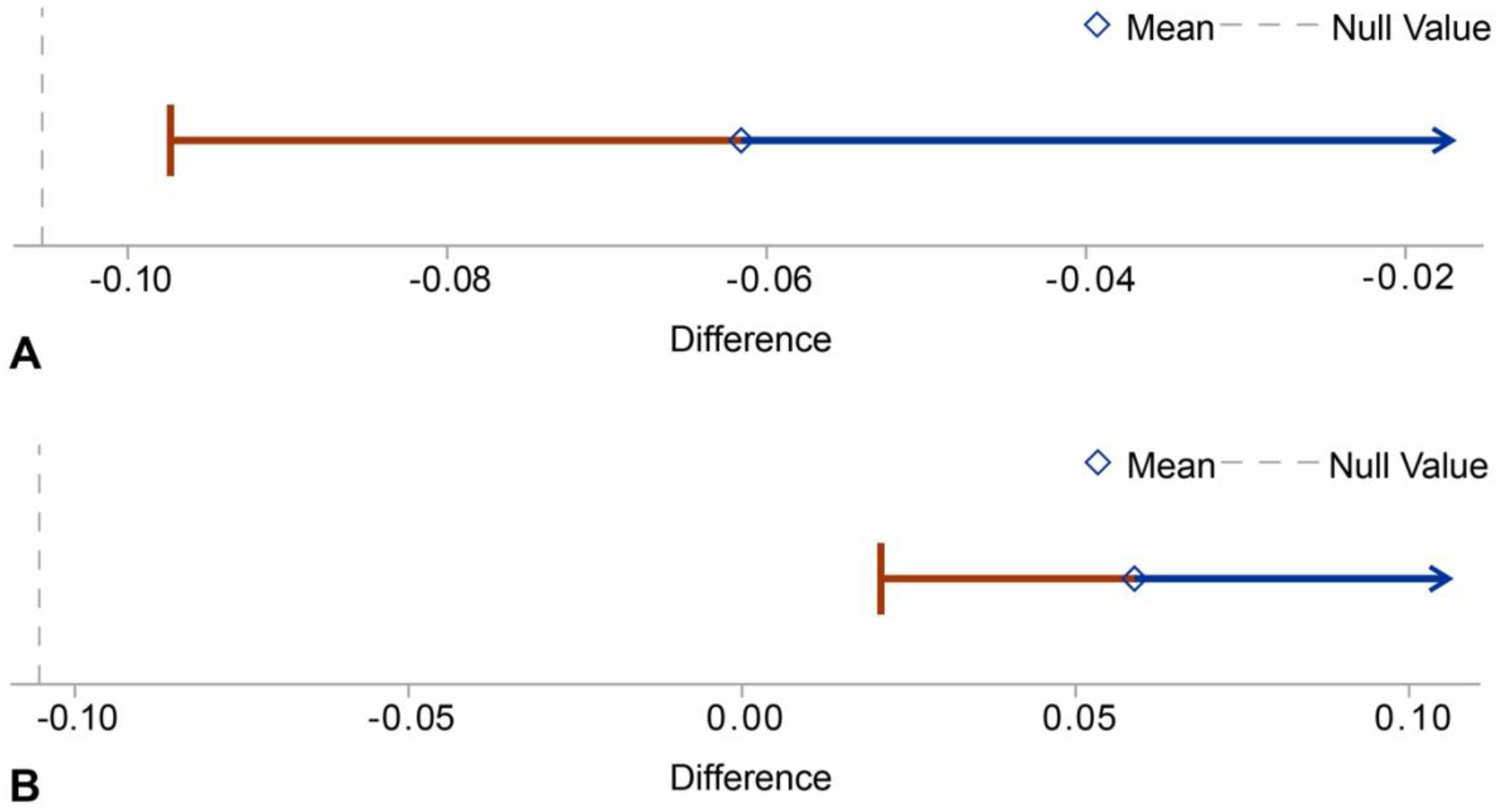
Results of the non-inferiority analysis of overall image quality for Rater A (A) and Rater B (B). The dashed vertical line on the far left indicates the non-inferiority margin (10%) obtained after logarithmic transformation of the mean differences between the two sequences. The diamond on the horizontal arrow represents the mean difference. Inferiority is rejected for both Rater A (*p* = 0.023) and Rater B (*p* < 0.0001), as the lower confidence limit (left-sided whisker of the one-sided 95% CI) lies above the non-inferiority margin.

**Figure 2.**
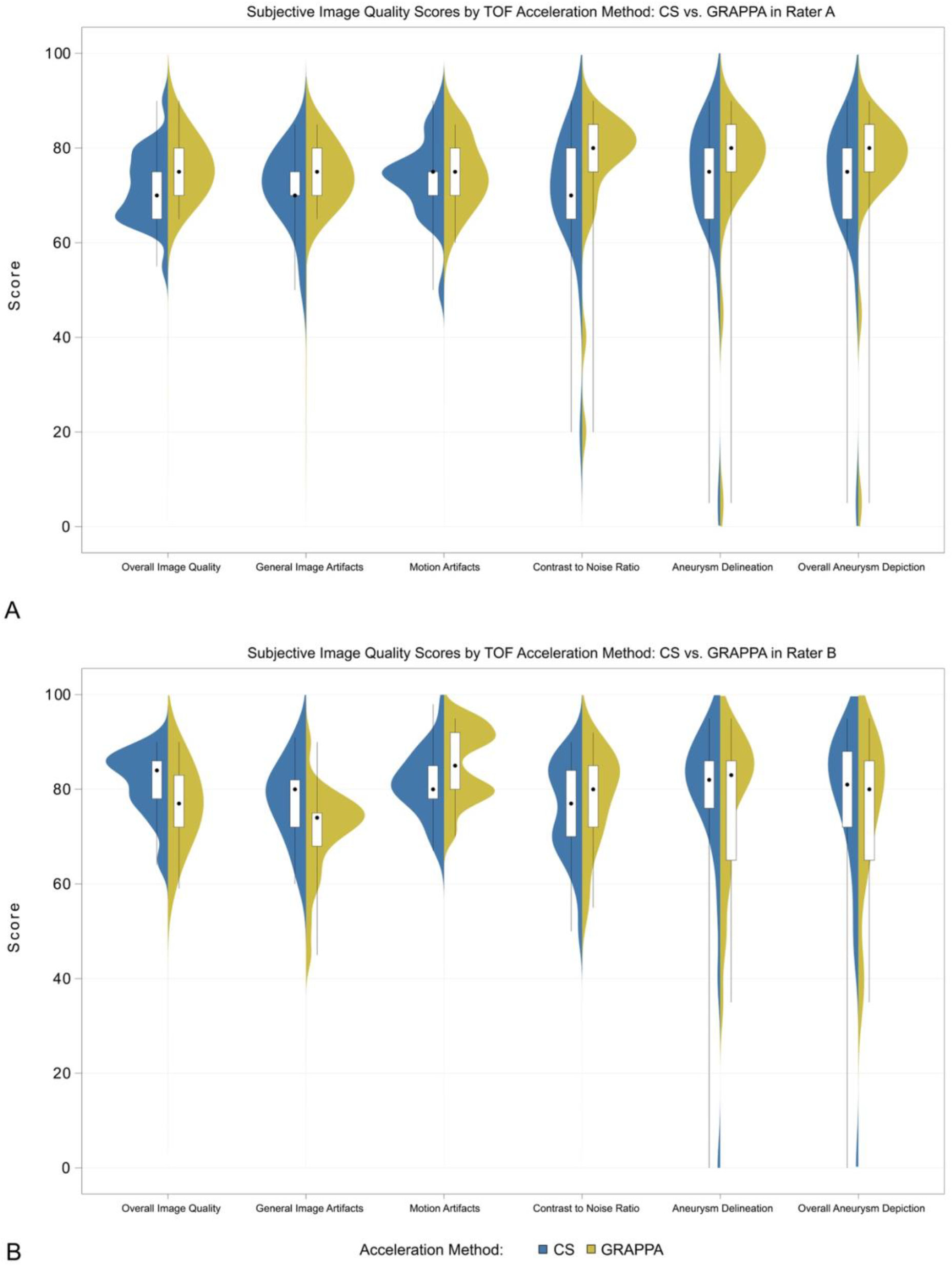
Subjective Image Quality Assessments by TOF acceleration Method in Rater A (A) and Rater B (B)

**Table 3.**
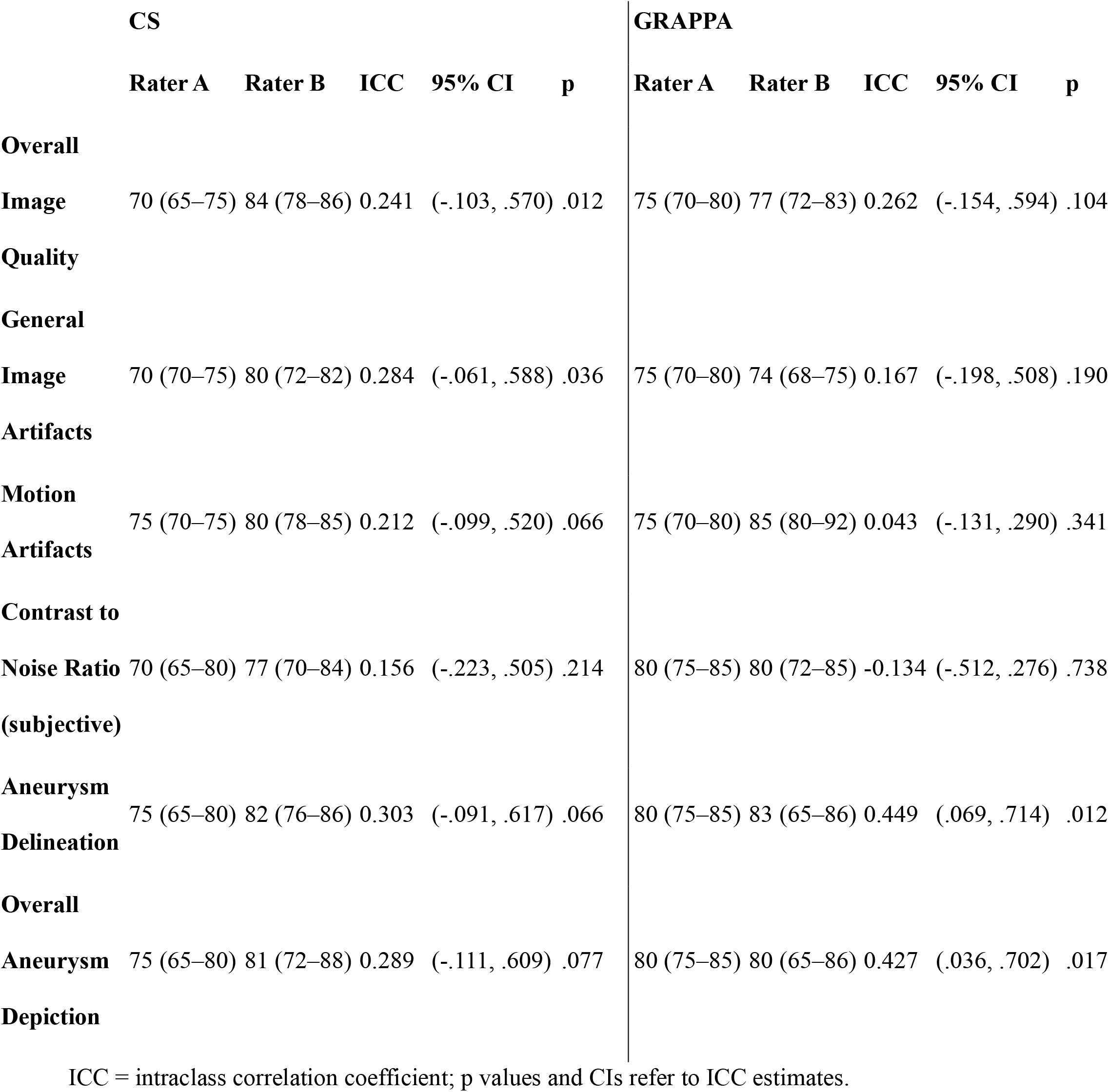
Subjective image quality ratings by sequence acceleration type and rater (0–100 scale).

### SECONDARY ENDPOINTS

Secondary subjective parameters, including general image artifacts, motion artifacts, subjective contrast-to-noise ratio, aneurysm delineation, and overall aneurysm depiction, were explored descriptively without formal non-inferiority testing (Table 3, Figure 2). Across these parameters, ratings demonstrated considerable interrater variability, with generally low to moderate interrater agreement. For several measures, Rater B tended to assign higher scores to CS-accelerated images, whereas Rater A more frequently favored GRAPPA-accelerated images. Median scores for most secondary endpoints were broadly comparable between acceleration methods, and no consistent pattern of superiority was observed across raters.

Objective contrast-to-noise ratio, obtained through ROI-based measurements, was higher for GRAPPA than for CS, with a median of 278.2 (IQR 181.0–366.2) compared with 169.6 (IQR 91.0–226.9), respectively (Figure 3).

**Figure 3.**
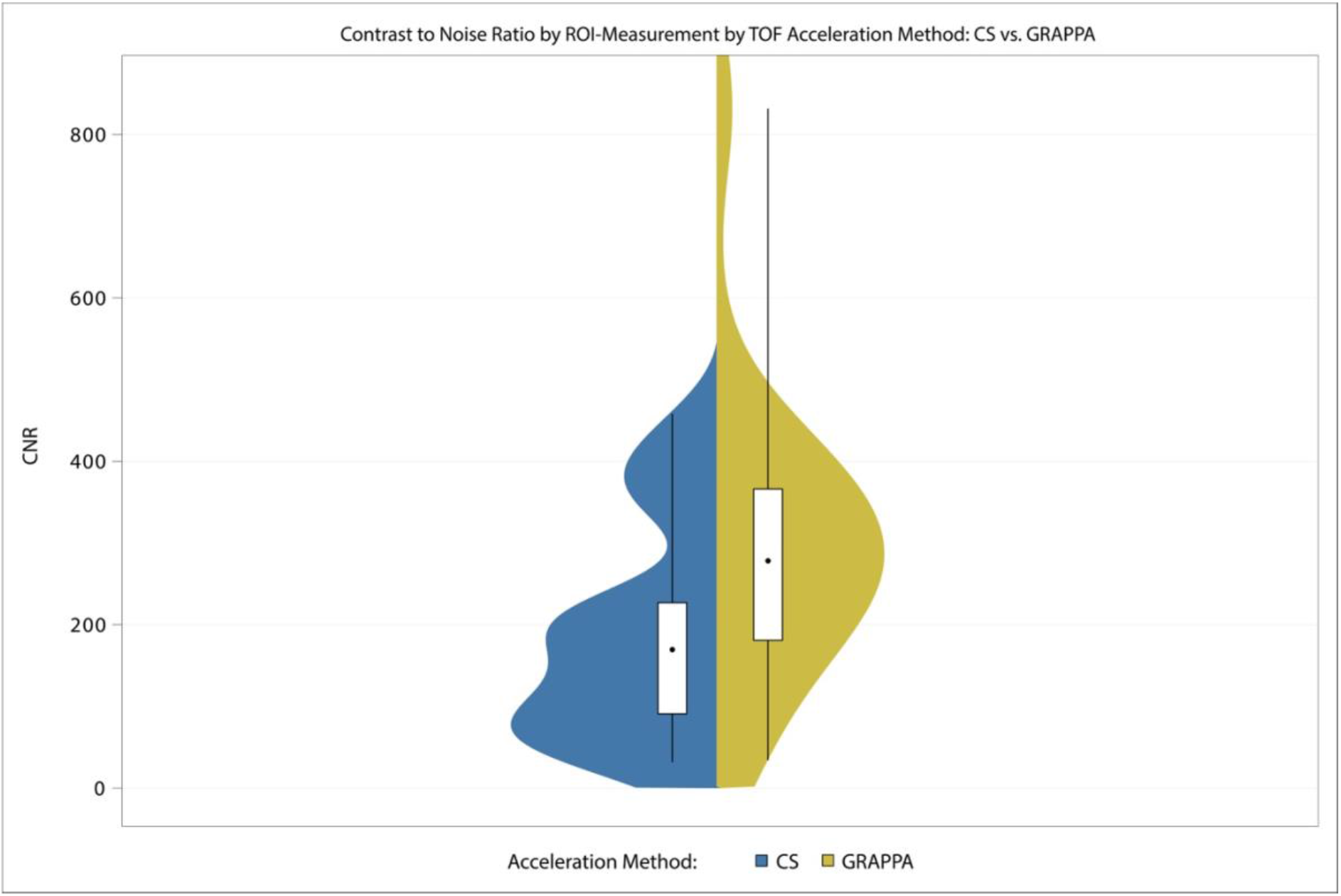
Distribution of ROI-based CNR ratios by acceleration method.

One submilimetric aneurysm arising from a dysplastic left middle cerebral artery (MCA), previously known from DSA, was not visualized by either reader with either CS- or GRAPPA-accelerated 3D-TOF. In another patient, Rater B identified a previously unknown extradural left ICA aneurysm measuring 3.4 mm in maximal diameter on GRAPPA-accelerated TOF-MRA, whereas the aneurysm was not detected by the same reader on CS-accelerated TOF-MRA. Rater A did not identify this aneurysm with either acceleration technique. In a further patient, Rater B did not identify on CS-accelerated TOF an aneurysm at the origin of the left anterior choroidal artery that measured up to 1.6 mm on the corresponding GRAPPA-accelerated TOF.

Measured aneurysm diameters demonstrated excellent interrater agreement across all dimensions and both acceleration methods (Table 4, Figure 4). For both CS- and GRAPPA-accelerated images, interrater agreement was consistently high for all measurements.

**Figure 4.**
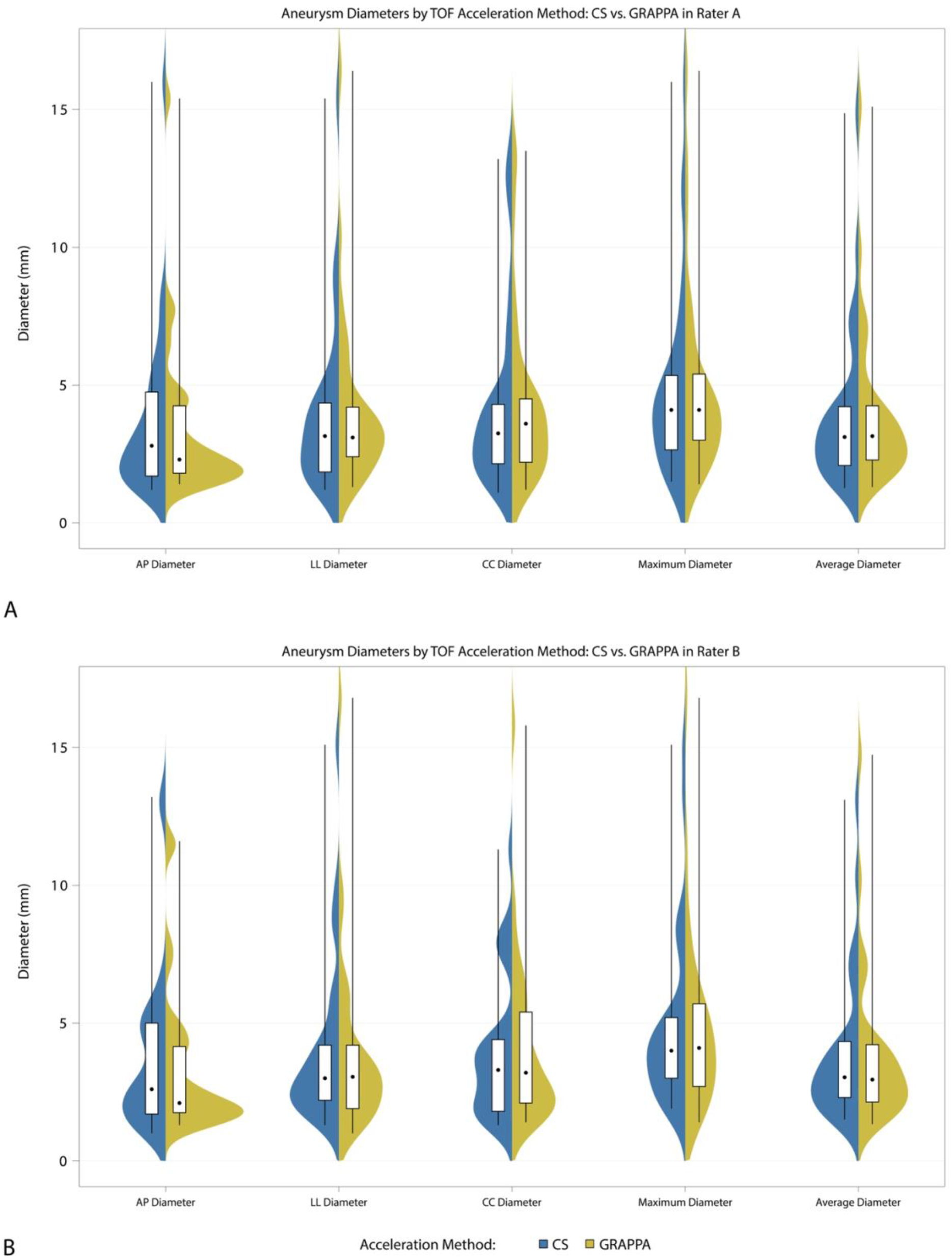
Distribution of measured aneurysm diameters by acceleration method and rater. AP = antero-posterior; LL = latero-lateral; CC = cranio-caudal.

**Figure 5.**
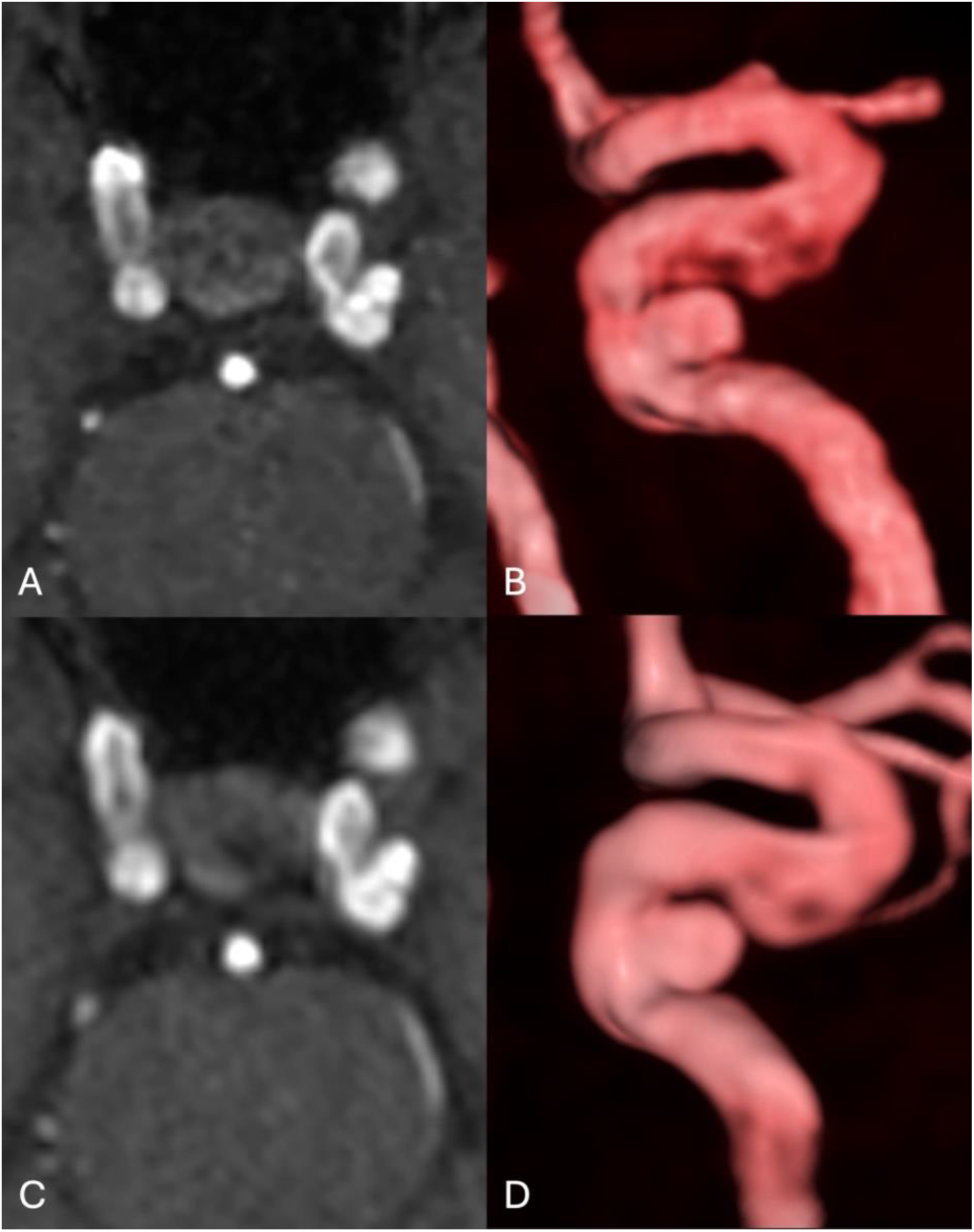
Axial CS-accelerated 3D-TOF-MRA (A) with corresponding 3D reconstruction (B), and axial GRAPPA-accelerated 3D-TOF-MRA (C) with corresponding 3D reconstruction (D), in a patient with a small left intracavernous infraophthalmic aneurysm. The aneurysm demonstrates non-inferior conspicuity on CS-accelerated TOF compared with GRAPPA-accelerated TOF, despite the lower CNR of the CS-accelerated acquisition.

**Table 4.**
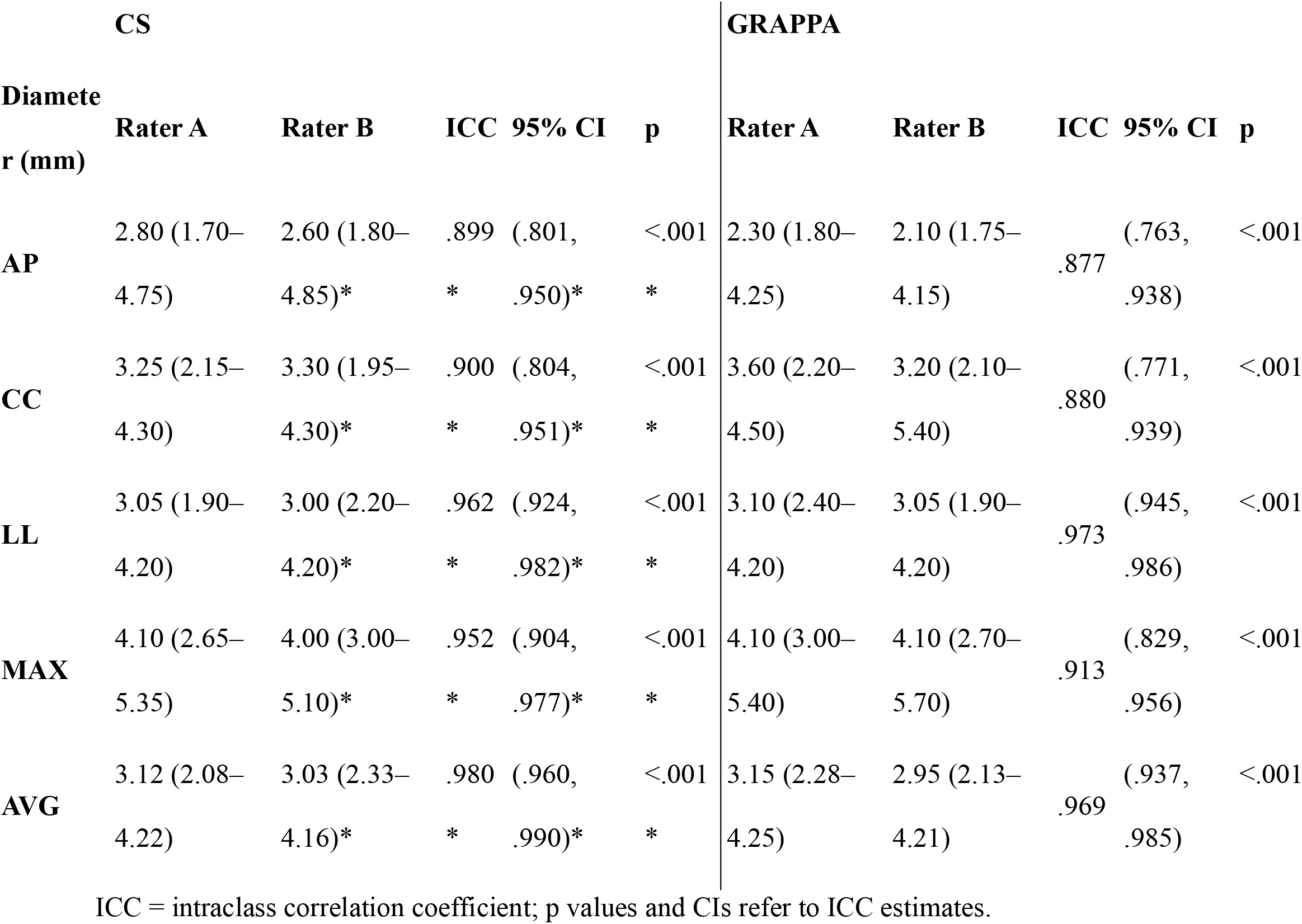
Measured aneurysm diameters by acceleration method and rater.

|  | **CS** |  |  |  |  | **GRAPPA** |  |  |  |  |
|---|---|---|---|---|---|---|---|---|---|---|
| **Diameter (mm)** | **Rater A** | **Rater B** | **ICC** | **95% CI** | **p** | **Rater A** | **Rater B** | **ICC** | **95% CI** | **p** |
| **AP** | 2.80 (1.70–4.75) | 2.60 (1.80–4.85)* | .899* | (.801, .950)* | <.001* | 2.30 (1.80–4.25) | 2.10 (1.75–4.15) | .877 | (.763, .938) | <.001 |
| **CC** | 3.25 (2.15–4.30) | 3.30 (1.95–4.30)* | .900* | (.804, .951)* | <.001* | 3.60 (2.20–4.50) | 3.20 (2.10–5.40) | .880 | (.771, .939) | <.001 |
| **LL** | 3.05 (1.90–4.20) | 3.00 (2.20–4.20)* | .962* | (.924, .982)* | <.001* | 3.10 (2.40–4.20) | 3.05 (1.90–4.20) | .973 | (.945, .986) | <.001 |
| **MAX** | 4.10 (2.65–5.35) | 4.00 (3.00–5.10)* | .952* | (.904, .977)* | <.001* | 4.10 (3.00–5.40) | 4.10 (2.70–5.70) | .913 | (.829, .956) | <.001 |
| **AVG** | 3.12 (2.08–4.22) | 3.03 (2.33–4.16)* | .980* | (.960, .990)* | <.001* | 3.15 (2.28–4.25) | 2.95 (2.13–4.21) | .969 | (.937, .985) | <.001 |

ICC = intraclass correlation coefficient; p values and CIs refer to ICC estimates.

Median aneurysm diameters and interquartile ranges were highly comparable between raters for both acceleration techniques across antero-posterior, cranio-caudal, latero-lateral, maximum, and average measurements. No systematic rater-or method-dependent bias was observed in any dimension.

In contrast to the low interrater agreement observed for subjective image quality assessments, these findings indicate that objective morphometric measurements are robust and reproducible and are largely unaffected by rater preference or acceleration technique.

## DISCUSSION

3D-TOF-MRA is an established method for evaluating intracranial aneurysms, offering high diagnostic performance without exposure to ionizing radiation or contrast agents (24).

However, long acquisition times, limited spatial resolution, and flow-related signal loss limit its clinical applicability. Acceleration strategies aim to shorten scan time and/or improve spatial resolution. PI techniques (GRAPPA, CAIPIRINHA) reduce acquisition time but incur SNR penalties at higher acceleration factors (5,8). In contrast, CS enables higher acceleration through incoherent k-space undersampling combined with iterative reconstruction (10,11).

TOF-MRA is particularly well suited to CS due to sparse vessel signal against a suppressed background.

In aneurysm focused studies, pretherapeutic 3T data indicates that CS-TOF is a suitable alternative, providing similar or superior image quality and diagnostic performance to PI-TOF, with equivalent aneurysm size measurements when validated against DSA (22,25). Additionally, CS-TOF offers equal or improved parent vessel visualization in coiled aneurysms (26).

Against this background, we prospectively compared two independently optimized CS- and GRAPPA-accelerated 3D-TOF-MRA sequences at 1.5T in patients with known untreated intracranial aneurysms. This extends existing evidence to a widely used clinical field strength in a controlled prospective setting.

In contrast to prior literature, our findings demonstrated marked heterogeneity in reader preference for CS-TOF versus GRAPPA-TOF, an observation limited however by the small number of readers (n=2). By contrast, Kim et al. reported a clear consensus favoring CS-TOF (p < 0.001), with substantial interobserver agreement (κ = 0.78–0.81) (22). Although each reader separately met the prespecified non-inferiority criterion, Rater A preferred GRAPPA (mean difference −0.062), whereas Rater B favored CS (mean difference 0.059), yielding low interrater agreement (ICC = 0.241–0.262) and opposite directional preferences. Potential explanations include field-strength-related image texture differences and the use of a 100- point continuous rating scale, which may detect subtle, reader-specific preferences not captured by coarser ordinal scales used in prior work.

This heterogeneity extended to exploratory secondary endpoints. Rater A consistently preferred GRAPPA across all evaluated parameters, while Rater B favored CS for overall image quality, general artifact burden, and aneurysm depiction, yet preferring GRAPPA for motion artifacts, subjective CNR, and aneurysm delineation. Subjective interpretation is known to be shaped by perceptual and cognitive factors, potentially accounting for limited interobserver agreement even in technically standardized datasets and indicating reader- specific weighting of features rather than systematic inferiority of either technique (27).

While these discrepancies were isolated, they might have disproportionately affected ratings and contributed to variability.

Outlier analyses further outline the sensitivity of subjective scoring to detection variability, as a few aneurysms showed discordant detection between techniques and readers. For instance, a single missed aneurysm affected multiple subjective metrics in Rater A, while in Rater B, a different aneurysm was not identified on CS but was detected on GRAPPA, and one additional aneurysm was identified exclusively on GRAPPA images. Similarly, a submillimetric MCA aneurysm was not visualized by either reader on either sequence, whereas a 3.4 mm extradural ICA aneurysm and a 1.6 mm anterior choroidal aneurysm were detected on GRAPPA but not CS-accelerated 3D-TOF by one reader. Although these discrepancies were isolated, they might have disproportionately affected ratings and contributed to variability. Aside from reader variability, these findings may also suggest reduced conspicuity of very small aneurysms when applying CS-acceleration, although the small number of discordant cases precludes definitive conclusions regarding differences in diagnostic sensitivity.

Quantitative measurements showed substantially lower CNR for CS than GRAPPA (CS median 169.6, IQR 91.0-226.9; GRAPPA median 278.2, IQR 181.0-366.2). This contrasts with several 3T reports where CS-TOF yielded higher CNR than PI-based accelerations, likely reflecting field-strength dependency: higher baseline SNR at 3T may allow CS to suppress noise while preserving signal, whereas 1.5T’s lower baseline signal may lead to disproportionate signal attenuation and reduced CNR (22,28).

ROI-based CNR comparisons between CS and GRAPPA are not straightforward due to distinct noise behaviours: CS uses incoherent undersampling with nonlinear iterative reconstruction, whereas GRAPPA relies on coil sensitivity information and is subject to spatially varying noise amplification (geometry factor). Moreover, CNR also depends on voxel size, nominal resolution, matrix and bandwidth, so lower CNR in CS-TOF should be interpreted cautiously as a sequence-specific trade-off rather than inherent inferiority (29).

Despite qualitative variability, aneurysm diameter measurements showed excellent interrater agreement for both methods (CS ICC 0.899–0.980; GRAPPA 0.877–0.973; all p < 0.001). This supports reproducible morphometric assessment, with CS not introducing systemic bias or clinically relevant geometric distortion under the parameters used (22).

Limitations include the lack of temporally matched DSA as a reference standard for most patients and the fact that readers were informed of the number and location of the aneurysms, precluding definitive assessment of diagnostic accuracy relative to ground truth. Accordingly, this study assessed equivalence rather than establish true sensitivity or specificity. Second, the opposing reader preferences and low interrater agreement slightly limit robustness and generalizability of the subjective non-inferiority conclusion. Third, the cohort was predominantly composed of small aneurysms (65.6% < 5 mm), thus limiting generalizability, particularly given evidence that 3D TOF-MRA has reduced sensitivity for aneurysms under 3 mm, may be affected by slow or turbulent flow in large or giant aneurysms and given our observation of reduced conspicuity in isolated very small aneurysms (30). Prior studies have suggested that PI may be preferred in the latter, potentially due to turbulence-related signal loss inherent to CS techniques (22). Further work is needed in complex morphologies and very small or larger aneurysms. Most aneurysms were in the anterior circulation and intradural, which limits extrapolation to posterior or extradural locations. Fourth, the optimized CS and GRAPPA protocols slightly differed in matrix and nominal spatial resolution. However, even at identical nominal resolution, a direct comparison would remain limited, as CS undersampling can reduce effective resolution despite higher nominal values.

Practically, CS provided a substantial acquisition-time reduction (38.7%) compared to GRAPPA. This aligns with prior reports, although acceleration gains vary with field strength and implemented acceleration factors (22,26,28). We used a relatively high acceleration factor (7) at 1.5T, which, despite maintaining non-inferior overall image quality coincided with a substantially reduced CNR and divergent reader preferences, suggesting that choice of acceleration factor may be limited at lower field strengths. Against the background of prior 3T studies, where CS acceleration generally benefited from higher intrinsic CNR, our findings indicate that the balance between acceleration, image quality, and perceptual consistency may be more constrained at 1.5T.

## CONCLUSION

At 1.5 T, the optimized CS-accelerated 3D-TOF sequence reduced acquisition time by 38.7% and met the prespecified 10% non-inferiority margin for overall image quality for both raters. However, opposing preferences, low interrater agreement and substantially lower ROI-based CNR demonstrate and important protocol trade-off. Aneurysm diameter measurements were highly reproducible between readers with both techniques. CS-accelerated 3D TOF-MRA at 1.5T is therefore as a time-efficient alternative to GRAPPA for evaluation of known intracranial aneurysms, although caution is warranted in very small aneurysms intracranial aneurysms, highlighting the need for further studies on field-strength dependency and optimal acceleration strategies.

## ACKNOWLEDGEMENTS

We acknowledge the contribution of Dipl.-Statistikerin Nadine Bonberg, who assisted with the estimation of the sample size required for the study. ChatGPT (OpenAI) was used to assist with language editing and to improve the readability of the manuscript. All scientific content, interpretations, and conclusions were generated, reviewed, and verified entirely by the authors.

## STATEMENTS AND DECLARATIONS

### ETHICAL CONSIDERATIONS

This single-center prospective non-inferiority study was approved by the institutional ethics committee of Carl von Ossietzky Universität, Oldenburg, Germany (No. 2021-033).

### CONSENT TO PARTICIPATE

All participants provided written informed consent, and procedures complied with the Declaration of Helsinki and applicable regulations.

### CONSENT FOR PUBLICATION

The authors affirm that human research participant provided informed consent for publication of the images in Figure 5.

### DECLARATION OF CONFLICTING INTEREST

Christian Mathys: consulting and lecturing for Siemens on behalf of the employer (Evangelisches Krankenhaus Oldenburg). The other authors declare that they have no known competing financial interests or personal relationships that could have appeared to influence the work reported in this paper.

### FUNDING STATEMENT

The project was funded through institutional resources. No external funding was received.

### DATA AVAILABILITY STATEMENT

The datasets generated and/or analyzed during the current study are not made publicly available due to local and European legal requirements governing the protection of patient data.

### AUTHORS’ CONTRIBUTIONS

Catalin George Iacoban, Christian Mathys and Benedikt Sundermann contributed to the initial study conception and design. Material preparation, data collection, and analysis were performed by Catalin George Iacoban, Holger Bösenberg, Claudia Klüner, Olga Alykova, Maike Gerdes, Franziska Meinert, Christian Mathys, and Benedikt Sundermann. The first draft of the manuscript was written by Catalin George Iacoban, and all authors reviewed and commented on previous versions of the manuscript. All authors have read and approved the final manuscript.

### PRESENTATION STATEMENT

The results of this study were presented in poster format at the 49th Annual Meeting of the European Society of Neuroradiology (ESNR), held jointly with the XXIII Symposium Neuroradiologicum and the British Society of Neuroradiologists (BSNR) Annual Meeting, in London, United Kingdom, from 30 September to 4 October 2026, are part of the doctoral thesis of the first author and will be submitted as a separate manuscript to a profile-specific medical journal.

## Notes

### Clinical Trial

https://drks.de/search/en/trial/DRKS00024403

### Author Declarations

This single-center prospective non-inferiority study was approved by the institutional ethics committee of Carl von Ossietzky University, Oldenburg, Germany (No. 2021-033).

