## supplementary material for "PROSPECTIVE COMPARISON OF OPTIMIZED COMPRESSED SENSING AND GRAPPA-ACCELERATED 3D-TOF-MRA FOR EVALUATION OF UNTREATED INTRACRANIAL ANEURYSMS AT 1.5 T: A NON-INFERIORITY STUDY"

### PRECURSOR PILOT STUDY FOR SAMPLE SIZE ESTIMATION

A retrospective pilot study was conducted to refine sample size estimation for the prospective phase. Fully anonymized 3D-TOF sequences comparable to the planned study protocol, acquired between January 1st and December 31st, 2021, were retrospectively retrieved from the PACS at the Institute of Radiology and Neuroradiology, Evangelisches Krankenhaus Oldenburg. The dataset included 20 chronologically consecutive routine MR examinations performed on the study scanner (1.5. T Magnetom Sola; SIEMENS Healthineers, Erlangen, Germany; Software version XA20, 20-channel head/neck receiver coil). Overall image quality was assessed independently by two blinded readers, followed by statistical analysis of data distributions. Due to lognormal data distribution of both datasets, the required sample size was calculated using a paired t-test for mean ratios. This method, available through SAS® Analytics Software, considers fixed parameters such as alpha error, nominal power, and inter-sequence correlations, while variations in results stem from differing coefficients of variation (CV). The sample-size calculation indicated 4 participants would be required based on the assessments of one reader and 8 participants based on those of the second reader. Using mean of these estimates (6 participants), the required sample-size increased to 7.2 subjects, after including the 20% drop-out reserve (the 20% potential data exclusions due to motion artifacts or incomplete examinations). Because the formally calculated sample size was considered unusually low, we adopted a more conservative approach and increased the planned sample size to 25 patients, consistent with sample sizes commonly used in comparable studies to enhance statistical power.

### SEQUENCE OPTIMIZATION

During the first stage of the prospective study, sequence parameters for CS (newer acceleration method) and for GRAPPA (conventional acceleration method) were optimized in five healthy participants using an approach similar to routine clinical protocol optimization by radiologists or MRI technologists. This pragmatic strategy relied on adjustments within predefined boundaries rather than exhaustive grid searches or simulation-based optimization.

To optimize and select the imaging sequences, multiple TOF acquisition were performed sequentially (Diagram 1), while scan times were maintained between 2.5 and 5 minutes and TR and TE were kept constant. Scan time reductions achieved with each technique were quantified. Sequence selection for further evaluation was based on consensus among experienced neuroradiologists (CM, BS, HB) and a general radiologist with 7 years' experience in neuroradiology (CGI). For each acceleration method arm (CS and GRAPPA), the two best-performing sequences were independently assessed (HB, CGI) based on the overall image quality (using an image rating reference catalogue) and ultimately the optimal sequence per arm was selected for use in the subsequent clinical phase.

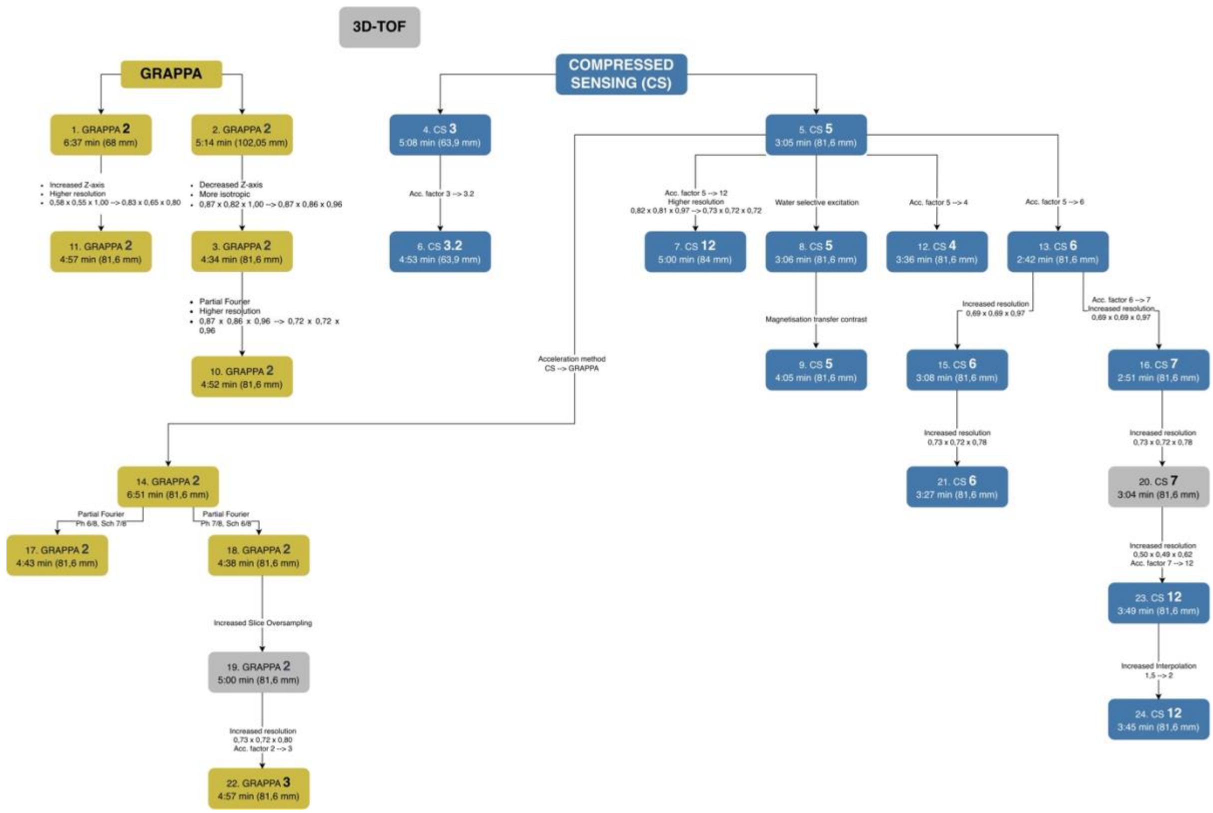


**Diagram S1.** Study workflow for the optimization and selection of 3D TOF sequence parameters in healthy volunteers. A total 24 GRAPPA- and CS-accelerated sequences were performed sequentially in healthy volunteers. Sequences 19 (GRAPPA) and 20 (CS), highlighted in grey, represent the final optimized sequences selected for the subsequent phase of the study.

**PATIENT COHORT**

During the sequence-optimization stage, 5 healthy volunteers with a minimum age of 18 were recruited. The final sample size for the aneurysm cohort was determined following completion of the retrospective pilot study and comprised 25 patients (including 20% drop-out rate) with at least one known untreated intracranial aneurysm. During the second prospective stage, from July 2024 to December 2024, 25 consecutive patients with at least one known untreated aneurysm (19 women; mean age, 61 years) underwent MR imaging as indicated by the treating physician, at the Institute of Radiology and Neuroradiology, Evangelisches Krankenhaus Oldenburg, using a 1.5 T MR scanner. General exclusion criteria included, but were not limited to: non-MR-conditional implants such as pacemakers or automatic defibrillators, implanted neural stimulators, ocular metallic foreign bodies, devices requiring a safety mode that poses increased risk to the patient (e.g., temporary deactivation of defibrillator function in MR-conditional devices), conditions that could impede study participation (e.g., morbid obesity, severe claustrophobia) and/or unnecessary or avoidable risks such as pregnancy, recent surgery of uncertain stability, the need for sedation, or other individual health factors or age-related concerns making MRI burdensome. Note: neurovascular implants (aneurysm clips, coils, stents, flow-diverters) were permitted if classified as MR-safe or MR-conditional, and if compliance with manufacturer-specified MRI safety parameters could be ensured and did not compromise the sequence parameter settings. Specific exclusion criteria for the study were known severe central nervous system disorders. Patients with additionally treated aneurysms (e.g., via clipping, coiling, stenting, or flow-diverter implantation) could be included, provided that these treatments did not constitute a contraindication to MRI.

**IMAGE ANALYSIS**

For each TOF acquisition, regions of interest (ROIs) were drawn on transversely reformatted slices to calculate the CNR within the following regions: 1. aneurysm lumen, 2. brain stem, and 3. background noise. The CNR was calculated as follows: CNR = (mean signal intensity of ANEURYSM LUMEN − mean signal intensity of BRAIN STEM) / (standard deviation of AIR).

Aneurysm diameters were measured in a standardized manner and recorded numerically in millimeters (mm). First, the maximum three-dimensional diameter was determined, independent of aneurysm neck location. Subsequently, the plane orthogonal to this maximum diameter was identified in which, based on visual estimation, the sum of the two largest perpendicular diameters appeared to be greatest. Within this plane, the two largest orthogonal diameters were then measured and recorded. Mean and maximum diameters were calculated. Comparison with digital subtraction angiography was not performed because previous DSA imaging was unavailable for most patients, which precluded reliable analysis.
